# A bibliometric analysis of the highest cited and highest category normalised articles in radiological literature from 2009-2019

**DOI:** 10.1101/2020.11.14.20231944

**Authors:** Nicholas McKay Parry, Justin Rich, Michael Erian, Thomas Lloyd

## Abstract

**Rationale and Objective:** Citation-based metrics are frequently used to evaluate the academic performance of a publication. One such metric is the number of citations an article receives, however this is not an infallible index. To account for biases of this metric the category-normalised citation index (CNCI), termed ‘impact’, may be used. Here the 100 highest-cited and highest-CNCI articles in radiological literature, from 2009 – 2019 is performed.

**Materials and Methods:** The Web of Science Core Collection and InCites (Clarivate Analytics) databases were accessed for the citations and CNCI values for articles published in the 186 journals with category “*radiology, nuclear medicine, and medical imaging*” between 2009 and 2019. The top 100 articles with the highest citation count and highest CNCI values were collected. Article parameters were analysed including title, year of publication, citation count, CNCI, field of study and modality studied were analysed.

**Results:** Fifty-three articles were common to both lists. Neuroradiology was the most prevalent subspecialty studied in both the highest-cited (n = 68) and highest-impact (n = 41) lists, respectively. The most frequently utilised imaging modality was magnetic resonance imaging (n = 64 and 40). The highest-CNCI articles demonstrated greater variability and distribution across subspecialties, imaging modalities and year of publication when compared to the highest-cited list.

**Conclusion:** The use of normalised bibliometric analysis tools may remove bias when evaluating research and better demonstrate the breadth of research activity. Use of these tools may provide a more robust and contemporaneous review of the landscape of research within a field.

## 1.0 Introduction

Bibliometric analysis is an established paradigm to compare the performance of academic output (1). One of the most common measures of performance is the raw citation count of an article - with the greater citation count reflecting greater significance. However, this metric fails to factor in the discipline of interest, age of publication and document type making it difficult to compare across academic disciplines (or sub-disciplines) as well as temporally and typically favours popularist topics. Other evaluative metrics exist to account for these biases and limitations, including the Category Normalised Citation Index (CNCI) (2,3). The CNCI is calculated by ratio of the actual count of citing items divided by the expected citation count for documents within the same field, year and type of article. Therefore, the CNCI *normalises* the citation count and expresses how much *impact* it has within its field. A CNCI of 2, for example, means the publication had twice as many citations as expected an article with a CNCI of 1. This creates a valuable measure of an articles significance which allows for comparison of articles in previous domains.

Bibliometric analyses is an increasingly popular method of defining significant reserch across medical literature (4–6). It serves to provide stakeholders (practitioners, researchers, trainees, etc) insights into the research. Citation and bibliometric analysis has been performed in the past within the radiological literature (7–11). However no studies, to the authors knowledge, has performed a category normalised citation index in radiological literature. By performing a bibliometric analysis with a normalised metric trends and impactful articles from less cited fields of research may emerge for interested stakeholders.

Our study aimed to determine the characteristics and trends of the 100 highest-cited and highest-CNCI’ articles in medical imaging literature between 2009 and 2019 within the Web of Science (WOS) database. In doing so both the highest-cited and highest-CNCI works in radiological journals are available to provide insight into contemporary radiological research. In addition this research serves to illustrate the utility of normalised bibliometrics.

## 2.0 Materials and Methods

In July 2020 the Web of Science (WOS) Core Collection (Clarivate Analytics) database was accessed to extract 186 journals with WOS category “Radiology, Nuclear Medicine & Medical Imaging” within the Science Citation Index Expanded (SCIE) and Emerging Source Citation Index (ESCI) respectively. The WOS database was limited to searches of these journals and the 100 articles with greatest number of citations between 2009 and 2019, inclusive, were recorded. Similarly, InCites (Clarivate Analytics) was accessed to extract the 100 articles with highest-CNCI. Articles excluded include those classified as primarily pertinent to radiation oncology and of the type “bibliographic, meeting article, preceding paper, news item, correction, letters to editor” were excluded due to biases in these article types which inflate their CNCI.

Each article within the highest-cited and highest-impact (CNCI) list were reviewed by two authors for categorisation. Following the template of Lim et al (10) and Brinkiki et al (11), data was collected on the following variables; (a) article demographics (journal name, impact factor of the journal, year of publication); (b) radiologic subspecialty (neuroradiology/head and neck [including studies related to fMRI], abdominal, breast, cardiac, respiratory, genitourinary [including retroperitoneum and obstetrics], musculoskeletal, paediatric, vascular/interventional, nuclear medicine, imaging safety or miscellaneous [not conforming to aforementioned categories including whole-body imaging, radiomics, physics/image processing and basic science]) and (c) radiographic technique/modality (conventional radiography, ultrasound, computed tomography (CT), magnetic resonance imaging (MRI), angiography, mammography, interventional, nuclear imaging [including PET, SPECT, PET/CT and PET/MRI], mixed and other [not conforming to above including image analysis tools]). Noting the radiologic subspeciality and radiographic modality are mutually exclusive variables.

Intersection over union calculations were performed to address similarity of articles via the Jaccard index (12). Analysis was performed using numerical computing environment MATLAB (R2018b; Mathworks, Massachusetts, USA).

## 3.0 Results

The 100 highest-cited works of 2009-2019 in radiological literature are presented in Table 1. The citation median was 694 (range 506 to 4008) and CNCI median 28.9 (range 9.31 to 198.8) and median year of publication 2011.

**Table 1.** Top 100 highest-citations articles in radiological literature (2009 – 2019).

| 100 Highest-Cited Articles in Radiology 2009-2019 |  |  |  |
| --- | --- | --- | --- |
| Rank | Article | Citations | CNCI |
| 1 | Rubinov, M. and Sporns, O., 2010. Complex network measures of brain connectivity: uses and interpretations. *Neuroimage*, 52(3), pp.1059-1069. | 4008 | 119.6 |
| 2 | Jenkinson, M., Beckmann, C.F., Behrens, T.E., Woolrich, M.W. and Smith, S.M., 2012. Fsl. *Neuroimage*, 62(2), pp.782-790. | 3327 | 60.8 |
| 3 | Power, J.D., Barnes, K.A., Snyder, A.Z., Schlaggar, B.L. and Petersen, S.E., 2012. Spurious but systematic correlations in functional connectivity MRI networks arise from subject motion. *Neuroimage*, 59(3), pp.2142-2154. | 3142 | 116.6 |
| 4 | Lang, R.M., Badano, L.P., Mor-Avi, V., Afilalo, J., Armstrong, A., Ernande, L., Flachskampf, F.A., Foster, E., Goldstein, S.A., Kuznetsova, T. and Lancellotti, P., 2015. Recommendations for cardiac chamber quantification by echocardiography in adults: an update from the American Society of Echocardiography and the European Association of Cardiovascular Imaging. *European Heart Journal-Cardiovascular Imaging*, 16(3), pp.233-271. | 2769 | 198.8 |
| 5 | Smith, S.M. and Nichols, T.E., 2009. Threshold-free cluster enhancement: addressing problems of smoothing, threshold dependence and localisation in cluster inference. *Neuroimage*, 44(1), pp.83-98. | 2375 | 64.9 |
| 6 | Fischl, B., 2012. FreeSurfer. *Neuroimage*, 62(2), pp.774-781. | 1797 | 32.8 |
| 7 | Klein, S., Staring, M., Murphy, K., Viergever, M.A. and Pluim, J.P., 2009. Elastix: a toolbox for intensity-based medical image registration. *IEEE transactions on medical imaging*, 29(1), pp.196-205. | 1547 | 59.5 |
| 8 | Nagueh, S.F., Smiseth, O.A., Appleton, C.P., Byrd, B.F., Dokainish, H., Edvardsen, T., Flachskampf, F.A., Gillebert, T.C., Klein, A.L., Lancellotti, P. and Marino, P., 2016. Recommendations for the evaluation of left ventricular diastolic function by echocardiography: an update from the American Society of Echocardiography and the European Association of Cardiovascular Imaging. *European Journal of Echocardiography*, 17(12), pp.1321-1360. | 1530 | 110.0 |
| 9 | Fedorov, A., Beichel, R., Kalpathy-Cramer, J., Finet, J., Fillion-Robin, J.C., Pujol, S., Bauer, C., Jennings, D., Fennessy, F., Sonka, M. and Buatti, J., 2012. 3D Slicer as an image computing platform for the Quantitative Imaging Network. *Magnetic resonance imaging*, 30(9), pp.1323-1341. | 1447 | 71.8 |
| 10 | Litjens, G., Kooi, T., Bejnordi, B.E., Setio, A.A.A., Ciompi, F., Ghafoorian, M., Van Der Laak, J.A., Van Ginneken, B. and Sánchez, C.I., 2017. A survey on deep learning in medical image analysis. *Medical image analysis*, 42, pp.60-88. | 1354 | 170.3 |
| 11 | Murphy, K., Birn, R.M., Handwerker, D.A., Jones, T.B. and Bandettini, P.A., 2009. The impact of global signal regression on resting state correlations: are anti-correlated networks introduced? *Neuroimage*, 44(3), pp.893-905. | 1353 | 37.0 |
| 12 | Van Dijk, K.R., Sabuncu, M.R. and Buckner, R.L., 2012. The influence of head motion on intrinsic functional connectivity MRI. *Neuroimage*, 59(1), pp.431-438. | 1351 | 50.1 |
| 13 | Nagueh, S.F., Appleton, C.P., Gillebert, T.C., Marino, P.N., Oh, J.K., Smiseth, O.A., Waggoner, A.D., Flachskampf, F.A., Pellikka, P.A. and Evangelisa, A., 2009. Recommendations for the evaluation of left ventricular diastolic function by echocardiography. *European Journal of Echocardiography*, 10(2), pp.165-193. | 1335 | 25.0 |
| 14 | Jacques, S.L., 2013. Optical properties of biological tissues: a review. *Physics in Medicine & Biology*, 58(11), p.R37. | 1317 | 37.2 |
| 15 | Barentsz, J.O., Richenberg, J., Clements, R., Choyke, P., Verma, S., Villeirs, G., Rouviere, O., Logager, V. and Fütterer, J.J., 2012. ESUR prostate MR guidelines 2012. *European radiology*, 22(4), pp.746-757. | 1282 | 63.7 |
| 16 | Klein, A., Andersson, J., Ardekani, B.A., Ashburner, J., Avants, B., Chiang, M.C., Christensen, G.E., Collins, D.L., Gee, J., Hellier, P. and Song, J.H., 2009. Evaluation of 14 nonlinear deformation algorithms applied to human brain MRI registration. *Neuroimage*, 46(3), pp.786-802. | 1188 | 32.5 |
| 17 | Van Essen, D.C., Smith, S.M., Barch, D.M., Behrens, T.E., Yacoub, E., Ugurbil, K. and Wu-Minn HCP Consortium, 2013. The WU-Minn human connectome project: an overview. *Neuroimage*, 80, pp.62-79. | 1168 | 51.3 |
| 18 | Gillies, R.J., Kinahan, P.E. and Hricak, H., 2016. Radiomics: images are more than pictures, they are data. *Radiology*, 278(2), pp.563-577. | 1152 | 120.5 |
| 19 | Woolrich, M.W., Jbabdi, S., Patenaude, B., Chappell, M., Makni, S., Behrens, T., Beckmann, C., Jenkinson, M. and Smith, S.M., 2009. Bayesian analysis of neuroimaging data in FSL. *Neuroimage*, 45(1), pp.S173-S186. | 1142 | 31.2 |
| 20 | Tustison, N.J., Avants, B.B., Cook, P.A., Zheng, Y., Egan, A., Yushkevich, P.A. and Gee, J.C., 2010. N4ITK: improved N3 bias correction. *IEEE transactions on medical imaging*, 29(6), pp.1310-1320. | 1131 | 43.5 |
| 21 | Avants, B.B., Tustison, N.J., Song, G., Cook, P.A., Klein, A. and Gee, J.C., 2011. A reproducible evaluation of ANTs similarity metric performance in brain image registration. *Neuroimage*, 54(3), pp.2033-2044. | 1130 | 38.1 |
| 22 | Winkler, A.M., Ridgway, G.R., Webster, M.A., Smith, S.M. and Nichols, T.E., 2014. Permutation inference for the general linear model. *Neuroimage*, 92, pp.381-397. | 1084 | 55.7 |
| 23 | Patenaude, B., Smith, S.M., Kennedy, D.N. and Jenkinson, M., 2011. A Bayesian model of shape and appearance for subcortical brain segmentation. *Neuroimage*, 56(3), pp.907-922. | 1065 | 35.9 |
| 24 | Jones, D.K., Knösche, T.R. and Turner, R., 2013. White matter integrity, fiber count, and other fallacies: the do's and don'ts of diffusion MRI. *Neuroimage*, 73, pp.239-254. | 1013 | 165.8 |
| 25 | Greve, D.N. and Fischl, B., 2009. Accurate and robust brain image alignment using boundary-based registration. *Neuroimage*, 48(1), pp.63-72. | 1002 | 27.4 |
| 26 | Stoodley, C.J. and Schmahmann, J.D., 2009. Functional topography in the human cerebellum: a meta-analysis of neuroimaging studies. *Neuroimage*, 44(2), pp.489-501. | 996 | 14.8 |
| 27 | Shin, H.C., Roth, H.R., Gao, M., Lu, L., Xu, Z., Noguees, I., Yao, J., Mollura, D. and Summers, R.M., 2016. Deep convolutional neural networks for computer-aided detection: CNN architectures, dataset characteristics and transfer learning. *IEEE transactions on medical imaging*, 35(5), pp.1285-1298. | 991 | 93.0 |
| 28 | Glasser, M.F., Sotiropoulos, S.N., Wilson, J.A., Coalson, T.S., Fischl, B., Andersson, J.L., Xu, J., Jbabdi, S., Webster, M., Polimeni, J.R. and Van Essen, D.C., 2013. The minimal preprocessing pipelines for the Human Connectome Project. *Neuroimage*, 80, pp.105-124. | 987 | 43.4 |
| 29 | Eickhoff, S.B., Laird, A.R., Grefkes, C., Wang, L.E., Zilles, K. and Fox, P.T., 2009. Coordinate-based activation likelihood estimation meta-analysis of neuroimaging data: A random-effects approach based on empirical estimates of spatial uncertainty. *Human brain mapping*, 30(9), pp.2907-2926. | 934 | 25.1 |
| 30 | Power, J.D., Mitra, A., Laumann, T.O., Snyder, A.Z., Schlaggar, B.L. and Petersen, S.E., 2014. Methods to detect, characterize, and remove motion artifact in resting state fMRI. *Neuroimage*, 84, pp.320-341. | 925 | 47.5 |
| 31 | Hutchison, R.M., Womelsdorf, T., Allen, E.A., Bandettini, P.A., Calhoun, V.D., Corbetta, M., Della Penna, S., Duyn, J.H., Glover, G.H., Gonzalez-Castillo, J. and Handwerker, D.A., 2013. Dynamic functional connectivity: promise, issues, and interpretations. *Neuroimage*, 80, pp.360-378. | 920 | 40.4 |
| 32 | Zalesky, A., Fornito, A. and Bullmore, E.T., 2010. Network-based statistic: identifying differences in brain networks. *Neuroimage*, 53(4), pp.1197-1207. | 892 | 25.1 |
| 33 | Destrieux, C., Fischl, B., Dale, A. and Halgren, E., 2010. Automatic parcellation of human cortical gyri and sulci using standard anatomical nomenclature. *Neuroimage*, 53(1), pp.1-15. | 884 | 25.0 |
| 34 | Price, C.J., 2012. A review and synthesis of the first 20 years of PET and fMRI studies of heard speech, spoken language and reading. *Neuroimage*, 62(2), pp.816-847. | 884 | 15.8 |
| 35 | Pereira, F., Mitchell, T. and Botvinick, M., 2009. Machine learning classifiers and fMRI: a tutorial overview. *Neuroimage*, 45(1), pp.S199-S209. | 878 | 23.5 |
| 36 | Smith, S.M., Miller, K.L., Salimi-Khorshidi, G., Webster, M., Beckmann, C.F., Nichols, T.E., Ramsey, J.D. and Woolrich, M.W., 2011. Network modelling methods for FMRI. *Neuroimage*, 54(2), pp.875-891. | 873 | 29.4 |
| 37 | Van Overwalle, F., 2009. Social cognition and the brain: a meta-analysis. *Human brain mapping*, 30(3), pp.829-858. | 869 | 12.9 |
| 38 | Lamm, C., Decety, J. and Singer, T., 2011. Meta-analytic evidence for common and distinct neural networks associated with directly experienced pain and empathy for pain. *Neuroimage*, 54(3), pp.2492-2502. | 857 | 28.9 |
| 39 | Reuter, M., Schmansky, N.J., Rosas, H.D. and Fischl, B., 2012. Within-subject template estimation for unbiased longitudinal image analysis. *Neuroimage*, 61(4), pp.1402-1418. | 851 | 31.6 |
| 40 | Zhang, H., Schneider, T., Wheeler-Kingshott, C.A. and Alexander, D.C., 2012. NODDI: practical in vivo neurite orientation dispersion and density imaging of the human brain. *Neuroimage*, 61(4), pp.1000-1016. | 831 | 30.8 |
| 41 | Christ, A., Kainz, W., Hahn, E.G., Honegger, K., Zefferer, M., Neufeld, E., Rascher, W., Janka, R., Bautz, W., Chen, J. and Kiefer, B., 2009. The Virtual Family—development of surface-based anatomical models of two adults and two children for dosimetric simulations. *Physics in Medicine & Biology*, 55(2), p.N23. | 767 | 28.0 |
| 42 | Leemans, A. and Jones, D.K., 2009. The B-matrix must be rotated when correcting for subject motion in DTI data. *Magnetic Resonance in Medicine: An Official Journal of the International Society for Magnetic Resonance in Medicine*, 61(6), pp.1336-1349. | 762 | 26.9 |
| 43 | Ferrari, M. and Quaresima, V., 2012. A brief review on the history of human functional near-infrared spectroscopy (fNIRS) development and fields of application. *Neuroimage*, 63(2), pp.921-935. | 748 | 13.1 |
| 44 | Lammer, J., Malagari, K., Vogl, T., Pilleul, F., Denys, A., Watkinson, A., Pitton, M., Sergeant, G., Pfammatter, T., Terraz, S. and Benhamou, Y., 2010. Prospective randomized study of doxorubicin-eluting-bead embolization in the treatment of hepatocellular carcinoma: results of the PRECISION V study. *Cardiovascular and interventional radiology*, 33(1), pp.41-52. | 737 | 27.9 |
| 45 | Boellaard, R., O'Doherty, M.J., Weber, W.A., Mottaghy, F.M., Lonsdale, M.N., Stroobants, S.G., Oyen, W.J., Kotzerke, J., Hoekstra, O.S., Pruim, J. and Marsden, P.K., 2010. FDG PET and PET/CT: EANM procedure guidelines for tumour PET imaging: version 1.0. *European journal of nuclear medicine and molecular imaging*, 37(1), p.181. | 730 | 29.2 |
| 46 | Stephan, K.E., Penny, W.D., Daunizeau, J., Moran, R.J. and Friston, K.J., 2009. Bayesian model selection for group studies. *Neuroimage*, 46(4), pp.1004-1017. | 708 | 18.9 |
| 47 | Menze, B.H., Jakab, A., Bauer, S., Kalpathy-Cramer, J., Farahani, K., Kirby, J., Burren, Y., Porz, N., Slotboom, J., Wiest, R. and Lanczi, L., 2014. The multimodal brain tumor image segmentation benchmark (BRATS). *IEEE transactions on medical imaging*, 34(10), pp.1993-2024. | 706 | 50.6 |
| 48 | Fonov, V., Evans, A.C., Botteron, K., Alml, C.R., McKinstry, R.C., Collins, D.L. and Brain Development Cooperative Group, 2011. Unbiased average age-appropriate atlases for pediatric studies. *Neuroimage*, 54(1), pp.313-327. | 704 | 23.1 |
| 49 | Yan, C.G., Cheung, B., Kelly, C., Colcombe, S., Craddock, R.C., Di Martino, A., Li, Q., Zuo, X.N., Castellanos, F.X. and Milham, M.P., 2013. A comprehensive assessment of regional variation in the impact of head micromovements on functional connectomics. *Neuroimage*, 76, pp.183-201. | 703 | 30.9 |
| 50 | Alsop, D.C., Detre, J.A., Golay, X., Günther, M., Hendrikse, J., Hernandez-Garcia, L., Lu, H., Macintosh, B.J., Parkes, L.M., Smits, M. and Van Osch, M.J., 2015. Recommended implementation of arterial spin-labeled perfusion MRI for clinical applications: a consensus of the ISMRM perfusion study group and the European consortium for ASL in dementia. *Magnetic resonance in medicine*, 73(1), pp.102-116. | 698 | 33.7 |
| 51 | Lancellotti, P., Moura, L., Pierard, L.A., Agricola, E., Popescu, B.A., Tribouilloy, C., Hagendorff, A., Monin, J.L., Badano, L., Zamorano, J.L. and European Association of Echocardiography, 2010. European Association of Echocardiography recommendations for the assessment of valvular regurgitation. Part 2: mitral and tricuspid regurgitation (native valve disease). *European Journal of Echocardiography*, 11(4), pp.307-332. | 690 | 126.3 |
| 52 | Heimann, T. and Meinzer, H.P., 2009. Statistical shape models for 3D medical image segmentation: a review. *Medical image analysis*, 13(4), pp.543-563. | 687 | 9.8 |
| 53 | Vercauteren, T., Pennec, X., Perchant, A. and Ayache, N., 2009. Diffeomorphic demons: Efficient non-parametric image registration. *NeuroImage*, 45(1), pp.S61-S72. | 687 | 18.4 |
| 54 | Boellaard, R., Delgado-Bolton, R., Oyen, W.J., Giammarile, F., Tatsch, K., Eschner, W., Verzijlbergen, F.J., Barrington, S.F., Pike, L.C., Weber, W.A. and Stroobants, S., 2015. FDG PET/CT: EANM procedure guidelines for tumour imaging: version 2.0. *European Journal of nuclear medicine and molecular imaging*, 42(2), pp.328-354. | 682 | 55.8 |
| 55 | Caspers, S., Zilles, K., Laird, A.R. and Eickhoff, S.B., 2010. ALE meta-analysis of action observation and imitation in the human brain. *Neuroimage*, 50(3), pp.1148-1167. | 670 | 19.5 |
| 56 | Chang, C. and Glover, G.H., 2010. Time-frequency dynamics of resting-state brain connectivity measured with fMRI. *Neuroimage*, 50(1), pp.81-98. | 667 | 19.4 |
| 57 | Van Essen, D.C., Ugurbil, K., Auerbach, E., Barch, D., Behrens, T.E.J., Bucholz, R., Chang, A., Chen, L., Corbetta, M., Curtiss, S.W. and Della Penna, S., 2012. The Human Connectome Project: a data acquisition perspective. *Neuroimage*, 62(4), pp.2222-2231. | 667 | 11.7 |
| 58 | Van Essen, D.C., Ugurbil, K., Auerbach, E., Barch, D., Behrens, T.E.J., Bucholz, R., Chang, A., Chen, L., Corbetta, M., Curtiss, S.W. and Della Penna, S., 2012. The Human Connectome Project: a data acquisition perspective. *Neuroimage*, 62(4), pp.2222-2231. | 664 | 26.0 |
| 59 | Van Overwalle, F. and Baetens, K., 2009. Understanding others' actions and goals by mirror and mentalizing systems: a meta-analysis. *Neuroimage*, 48(3), pp.564-584. | 663 | 9.8 |
| 60 | Rengier, F., Mehndiratta, A., Von Tengg-Kobligh, H., Zechmann, C.M., Unterhinninghofen, R., Kauczor, H.U. and Giesel, F.L., 2010. 3D printing based on imaging data: review of medical applications. *International journal of computer assisted radiology and surgery*, 5(4), pp.335-341. | 663 | 14.7 |
| 61 | Hampshire, A., Chamberlain, S.R., Monti, M.M., Duncan, J. and Owen, A.M., 2010. The role of the right inferior frontal gyrus: inhibition and attentional control. *Neuroimage*, 50(3), pp.1313-1319. | 656 | 19.1 |
| 62 | Satterthwaite, T.D., Elliott, M.A., Gerraty, R.T., Ruparel, K., Loughhead, J., Calkins, M.E., Eickhoff, S.B., Hakonarson, H., Gur, R.C., Gur, R.E. and Wolf, D.H., 2013. An improved framework for confound regression and filtering for control of motion artifact in the preprocessing of resting-state functional connectivity data. *Neuroimage*, 64, pp.240-256. | 645 | 28.4 |
| 63 | Treeby, B.E. and Cox, B.T., 2010. k-Wave: MATLAB toolbox for the simulation and reconstruction of photoacoustic wave fields. *Journal of biomedical optics*, 15(2), p.021314. | 639 | 24.8 |
| 64 | Craddock, R.C., James, G.A., Holtzheimer III, P.E., Hu, X.P. and Mayberg, H.S., 2012. A whole brain fMRI atlas generated via spatially constrained spectral clustering. *Human brain mapping*, 33(8), pp.1914-1928. | 637 | 22.9 |
| 65 | Bartra, O., McGuire, J.T. and Kable, J.W., 2013. The valuation system: a coordinate-based meta-analysis of BOLD fMRI experiments examining neural correlates of subjective value. *Neuroimage*, 76, pp.412-427. | 637 | 28.0 |
| 66 | Zalesky, A., Fornito, A., Harding, I.H., Cocchi, L., Yücel, M., Pantelis, C. and Bullmore, E.T., 2010. Whole-brain anatomical networks: does the choice of nodes matter?. *Neuroimage*, 50(3), pp.970-983. | 631 | 18.4 |
| 67 | Marks, L.B., Yorke, E.D., Jackson, A., Ten Haken, R.K., Constine, L.S., Eisbruch, A., Bentzen, S.M., Nam, J. and Deasy, J.O., 2010. Use of normal tissue complication probability models in the clinic. *International Journal of Radiation Oncology\* Biology\* Physics*, 76(3), pp.S10-S19. | 619 | 20.4 |
| 68 | McDonald, R.J., McDonald, J.S., Kallmes, D.F., Jentoft, M.E., Murray, D.L., Thielen, K.R., Williamson, E.E. and Eckel, L.J., 2015. Intracranial gadolinium deposition after contrast-enhanced MR imaging. *Radiology*, 275(3), pp.772-782. | 619 | 50.7 |
| 69 | McLaren, D.G., Ries, M.L., Xu, G. and Johnson, S.C., 2012. A generalized form of context-dependent psychophysiological interactions (gPPI): a comparison to standard approaches. *Neuroimage*, 61(4), pp.1277-1286. | 612 | 22.0 |
| 70 | Zuo, X.N., Di Martino, A., Kelly, C., Shehzad, Z.E., Gee, D.G., Klein, D.F., Castellanos, F.X., Biswal, B.B. and Milham, M.P., 2010. The oscillating brain: complex and reliable. *Neuroimage*, 49(2), pp.1432-1445. | 604 | 19.9 |
| 71 | Setsompop, K., Gagoski, B.A., Polimeni, J.R., Witzel, T., Wedeen, V.J. and Wald, L.L., 2012. Blipped-controlled aliasing in parallel imaging for simultaneous multislice echo planar imaging with reduced g-factor penalty. *Magnetic resonance in medicine*, 67(5), pp.1210-1224. | 600 | 28.9 |
| 72 | Zhang, D., Wang, Y., Zhou, L., Yuan, H., Shen, D. and Alzheimer's Disease Neuroimaging Initiative, 2011. Multimodal classification of Alzheimer's disease and mild cognitive impairment. *Neuroimage*, 55(3), pp.856-867. | 597 | 19.6 |
| 73 | Wheeler-Kingshott, C.A. and Cercignani, M., 2009. About "axial" and "radial" diffusivities. *Magnetic Resonance in Medicine: An Official Journal of the International Society for Magnetic Resonance in Medicine*, 61(5), pp.1255-1260. | 595 | 19.8 |
| 74 | Uddin, L.Q., Clare Kelly, A.M., Biswal, B.B., Xavier Castellanos, F. and Milham, M.P., 2009. Functional connectivity of default mode network components: correlation, anticorrelation, and causality. *Human brain mapping*, 30(2), pp.625-637. | 592 | 15.8 |
| 75 | Van Den Heuvel, M.P., Mandl, R.C., Kahn, R.S. and Hulshoff Pol, H.E., 2009. Functionally linked resting-state networks reflect the underlying structural connectivity architecture of the human brain. *Human brain mapping*, 30(10), pp.3127-3141. | 587 | 15.7 |
| 76 | Lu, G. and Fei, B., 2014. Medical hyperspectral imaging: a review. *Journal of biomedical optics*, 19(1), p.010901. | 586 | 15.8 |
| 77 | Eickhoff, S.B., Bzdok, D., Laird, A.R., Kurth, F. and Fox, P.T., 2012. Activation likelihood estimation meta-analysis revisited. *Neuroimage*, 59(3), pp.2349-2361. | 584 | 21.0 |
| 78 | International Commission on Non-Ionizing Radiation Protection, 2010. Guidelines for limiting exposure to time-varying electric and magnetic fields (1 Hz to 100 kHz). *Health physics*, 99(6), pp.818-836. | 582 | 30.2 |
| 79 | Kamnitsas, K., Ledig, C., Newcombe, V.F., Simpson, J.P., Kane, A.D., Menon, D.K., Rueckert, D. and Glocker, B., 2017. Efficient multi-scale 3D CNN with fully connected CRF for accurate brain lesion segmentation. *Medical image analysis*, 36, pp.61-78. | 579 | 72.8 |
| 80 | Satterthwaite, T.D., Wolf, D.H., Loughhead, J., Ruparel, K., Elliott, M.A., Hakonarson, H., Gur, R.C. and Gur, R.E., 2012. Impact of in-scanner head motion on multiple measures of functional connectivity: relevance for studies of neurodevelopment in youth. *Neuroimage*, 60(1), pp.623-632. | 578 | 20.8 |
| 81 | Jensen, J.H. and Helpert, J.A., 2010. MRI quantification of non-Gaussian water diffusion by kurtosis analysis. *NMR in Biomedicine*, 23(7), pp.698-710. | 577 | 11.1 |
| 82 | Armato III, S.G., McLennan, G., Bidaut, L., McNitt-Gray, M.F., Meyer, C.R., Reeves, A.P., Zhao, B., Aberle, D.R., Henschke, C.I., Hoffman, E.A. and Kazerooni, E.A., 2011. The lung image database consortium (LIDC) and image database resource initiative (IDRI): a completed reference database of lung nodules on CT scans. *Medical physics*, 38(2), pp.915-931. | 570 | 22.2 |
| 83 | Havaei, M., Davy, A., Warde-Farley, D., Biard, A., Courville, A., Bengio, Y., Pal, C., Jodoin, P.M. and Larochelle, H., 2017. Brain tumor segmentation with deep neural networks. *Medical image analysis*, 35, pp.18-31. | 570 | 71.7 |
| 84 | Winkler, A.M., Kochunov, P., Blangero, J., Almasy, L., Zilles, K., Fox, P.T., Duggirala, R. and Glahn, D.C., 2010. Cortical thickness or grey matter volume? The importance of selecting the phenotype for imaging genetics studies. *Neuroimage*, 53(3), pp.1135-1146. | 560 | 16.3 |
| 85 | Spreng, R.N., Stevens, W.D., Chamberlain, J.P., Gilmore, A.W. and Schacter, D.L., 2010. Default network activity, coupled with the frontoparietal control network, supports goal-directed cognition. *Neuroimage*, 53(1), pp.303-317. | 558 | 16.2 |
| 86 | Kumar, V., Gu, Y., Basu, S., Berglund, A., Eschrich, S.A., Schabath, M.B., Forster, K., Aerts, H.J., Dekker, A., Fenstermacher, D. and Goldof, D.B., 2012. Radiomics: the process and the challenges. *Magnetic resonance imaging*, 30(9), pp.1234-1248. | 557 | 27.7 |
| 87 | Tajbakhsh, N., Shin, J.Y., Gurudu, S.R., Hurst, R.T., Kendall, C.B., Gotway, M.B. and Liang, J., 2016. Convolutional neural networks for medical image analysis: Full training or fine tuning?. *IEEE transactions on medical imaging*, 35(5), pp.1299-1312. | 557 | 52.3 |
| 88 | Calhoun, V.D., Liu, J. and Adali, T., 2009. A review of group ICA for fMRI data and ICA for joint inference of imaging, genetic, and ERP data. *Neuroimage*, 45(1), pp.S163-S172. | 554 | 14.8 |
| 89 | Blankertz, B., Lemm, S., Treder, M., Haufe, S. and Müller, K.R., 2011. Single-trial analysis and classification of ERP components—a tutorial. *NeuroImage*, 56(2), pp.814-825. | 552 | 18.1 |
| 90 | Sotiras, A., Davatzikos, C. and Paragios, N., 2013. Deformable medical image registration: A survey. *IEEE transactions on medical imaging*, 32(7), pp.1153-1190. | 552 | 29.9 |
| 91 | Haacke, E.M., Mittal, S., Wu, Z., Neelavalli, J. and Cheng, Y.C., 2009. Susceptibility-weighted imaging: technical aspects and clinical applications, part 1. *American Journal of Neuroradiology*, 30(1), pp.19-30. | 548 | 9.3 |
| 92 | Aljabar, P., Heckemann, R.A., Hammers, A., Hajnal, J.V. and Rueckert, D., 2009. Multi-atlas based segmentation of brain images: atlas selection and its effect on accuracy. *Neuroimage*, 46(3), pp.726-738. | 547 | 14.6 |
| 93 | Kanda, T., Ishii, K., Kawaguchi, H., Kitajima, K. and Takenaka, D., 2014. High signal intensity in the dentate nucleus and globus pallidus on unenhanced T1-weighted MR images: relationship with increasing cumulative dose of a gadolinium-based contrast material. *Radiology*, 270(3), pp.834-841. | 541 | 36.3 |
| 94 | Eiber, 2016. " Evaluation of Hybrid Ga-68-PSMA Ligand PET/CT in 248 Patients with Biochemical Recurrence After Radical Prostatectomy"(vol 56, pg 668, 2015). *Journal of nuclear medicine*, 57(8), pp.1325-1325. | 538 | 44.0 |
| 95 | Andersson, J.L. and Sotiropoulos, S.N., 2016. An integrated approach to correction for off-resonance effects and subject movement in diffusion MR imaging. *Neuroimage*, 125, pp.1063-1078. | 538 | 45.6 |
| 96 | Sodickson, A., Baeyens, P.F., Andriole, K.P., Prevedello, L.M., Nawfel, R.D., Hanson, R. and Khorasani, R., 2009. Recurrent CT, cumulative radiation exposure, and associated radiation-induced cancer risks from CT of adults. *Radiology*, 251(1), pp.175-184. | 534 | 18.9 |
| 97 | Clark, K., Vendt, B., Smith, K., Freymann, J., Kirby, J., Koppel, P., Moore, S., Phillips, S., Maffitt, D., Pringle, M. and Tarbox, L., 2013. The Cancer Imaging Archive (TCIA): maintaining and operating a public information repository. *Journal of digital imaging*, 26(6), pp.1045-1057. | 527 | 29.7 |
| 98 | Bamber, J., Cosgrove, D., Dietrich, C.F., Fromageau, J., Bojunga, J., Calliada, F., Cantisani, V., Correas, J.M., D'onofrio, M., Drakonaki, E.E. and Fink, M., 2013. EFSUMB guidelines and recommendations on the clinical use of ultrasound elastography. Part 1: Basic principles and technology. *Ultraschall in der Medizin-European Journal of Ultrasound*, 34(02), pp.169-184. | 523 | 33.6 |
| 99 | Piscaglia, F., Nolsøe, C., Dietrich, C.A., Cosgrove, D.O., Gilja, O.H., Nielsen, M.B., Albrecht, T., Barozzi, L., Bertolotto, M., Catalano, O. and Claudon, M., 2012. The EFSUMB Guidelines and Recommendations on the Clinical Practice of Contrast Enhanced Ultrasound (CEUS): update 2011 on non-hepatic applications. *Ultraschall in der Medizin-European Journal of Ultrasound*, 33(01), pp.33-59. | 506 | 29.3 |
| 100 | Afshar-Oromieh, A., Zechmann, C.M., Malcher, A., Eder, M., Eisenhut, M., Linhart, H.G., Holland-Letz, T., Hadaschik, B.A., Giesel, F.L., Debus, J. and Haberkorn, U., 2014. Comparison of PET imaging with a 68 Ga-labelled PSMA ligand and 18 F-choline-based PET/CT for the diagnosis of recurrent prostate cancer. *European journal of nuclear medicine and molecular imaging*, 41(1), pp.11-20. | 506 | 33.9 |

The 100 highest-CNCI works between 2009-2019 are presented in Table 2. The citation median was 533 (range 42 to 4008) and CNCI median was 37.9 (range 27 to 198.8), and median year of publication 2013. Figure 1 presents a box and whisker plot to compare the distribution of articles between the two lists in terms of their raw citation count and CNCI. Table 3 presents the number of articles within each list published for a given year. Fifty-three articles were common to both lists with a Jaccard similarity index of 36%, demonstrating significant differences (low overlap) between the elements of each list.

**Table 2.** Top 100 highest-impact articles in radiological literature (2009 – 2019).

| 100 Highest-CNCI Articles in Radiology (2009-2019) |  |  |  |
| --- | --- | --- | --- |
| Rank | Article | Citations | CNCI |
| 1 | Lang, R.M., Badano, L.P., Mor-Avi, V., Afilalo, J., Armstrong, A., Ernande, L., Flachskampf, F.A., Foster, E., Goldstein, S.A., Kuznetsova, T. and Lancellotti, P., 2015. Recommendations for cardiac chamber quantification by echocardiography in adults: an update from the American Society of Echocardiography and the European Association of Cardiovascular Imaging. *European Heart Journal-Cardiovascular Imaging*, 16(3), pp.233-271. | 2769 | 198.8 |
| 2 | Litjens, G., Kooi, T., Bejnordi, B.E., Setio, A.A.A., Ciompi, F., Ghafoorian, M., Van Der Laak, J.A., Van Ginneken, B. and Sánchez, C.I., 2017. A survey on deep learning in medical image analysis. *Medical image analysis*, 42, pp.60-88. | 1354 | 170.3 |
| 3 | Jones, D.K., Knösche, T.R. and Turner, R., 2013. White matter integrity, fiber count, and other fallacies: the do's and don'ts of diffusion MRI. *Neuroimage*, 73, pp.239-254. | 1013 | 165.8 |
| 4 | Greenspan, H., Van Ginneken, B. and Summers, R.M., 2016. Guest editorial deep learning in medical imaging: Overview and future promise of an exciting new technique. *IEEE Transactions on Medical Imaging*, 35(5), pp.1153-1159. | 424 | 139.9 |
| 5 | Oliveira Melo, A.S., Malinger, G., Ximenes, R., Szejnfeld, P.O., Alves Sampaio, S. and Bispo de Filippis, A.M., 2016. Zika virus intrauterine infection causes fetal brain abnormality and microcephaly: tip of the iceberg?. *Ultrasound in Obstetrics & Gynecology*, 47(1), pp.6-7. | 409 | 136.7 |
| 6 | Gillies, R.J., Kinahan, P.E. and Hricak, H., 2016. Radiomics: images are more than pictures, they are data. *Radiology*, 278(2), pp.563-577. | 1152 | 120.5 |
| 7 | Rubinov, M. and Sporns, O., 2010. Complex network measures of brain connectivity: uses and interpretations. *Neuroimage*, 52(3), pp.1059-1069. | 4008 | 119.6 |
| 8 | Power, J.D., Barnes, K.A., Snyder, A.Z., Schlaggar, B.L. and Petersen, S.E., 2012. Spurious but systematic correlations in functional connectivity MRI networks arise from subject motion. *Neuroimage*, 59(3), pp.2142-2154. | 3142 | 116.6 |
| 9 | Shin, H.C., Roth, H.R., Gao, M., Lu, L., Xu, Z., Nogues, I., Yao, J., Mollura, D. and Summers, R.M., 2016. Deep convolutional neural networks for computer-aided detection: CNN architectures, dataset characteristics and transfer learning. *IEEE transactions on medical imaging*, 35(5), pp.1285-1298. | 991 | 93.0 |
| 10 | Kamnitsas, K., Ledig, C., Newcombe, V.F., Simpson, J.P., Kane, A.D., Menon, D.K., Rueckert, D. and Glocker, B., 2017. Efficient multi-scale 3D CNN with fully connected CRF for accurate brain lesion segmentation. *Medical image analysis*, 36, pp.61-78. | 579 | 72.8 |
| 11 | Fedorov, A., Beichel, R., Kalpathy-Cramer, J., Finet, J., Fillion-Robin, J.C., Pujol, S., Bauer, C., Jennings, D., Fennessy, F., Sonka, M. and Buatti, J., 2012. 3D Slicer as an image computing platform for the Quantitative Imaging Network. *Magnetic resonance imaging*, 30(9), pp.1323-1341. | 1447 | 71.8 |
| 12 | Havaei, M., Davy, A., Warde-Farley, D., Biard, A., Courville, A., Bengio, Y., Pal, C., Jodoin, P.M. and Larochelle, H., 2017. Brain tumor segmentation with deep neural networks. *Medical image analysis*, 35, pp.18-31. | 570 | 71.7 |
| 13 | Smith, S.M. and Nichols, T.E., 2009. Threshold-free cluster enhancement: addressing problems of smoothing, threshold dependence and localisation in cluster inference. *Neuroimage*, 44(1), pp.83-98. | 2375 | 64.9 |
| 14 | Barentsz, J.O., Richenberg, J., Clements, R., Choyke, P., Verma, S., Villeirs, G., Rouviere, O., Logager, V. and Fütterer, J.J., 2012. ESUR prostate MR guidelines 2012. *European radiology*, 22(4), pp.746-757. | 1282 | 63.7 |
| 15 | Chen, J., Lemyre, L., Wilkins, R. and Wilkinson, D., 2010. Radiation Protection Dosimetry. Preface. *Radiation protection dosimetry*, 142(1), pp.1-1. | 311 | 62.9 |
| 16 | Wahl, R.L., Jacene, H., Kasamon, Y. and Lodge, M.A., 2009. From RECIST to PERCIST: evolving considerations for PET response criteria in solid tumors. *Journal of nuclear medicine: official publication, Society of Nuclear Medicine*, 50(Suppl 1), p.122S. | 1708 | 61.6 |
| 17 | Jenkinson, M., Beckmann, C.F., Behrens, T.E., Woolrich, M.W. and Smith, S.M., 2012. Fsl. *Neuroimage*, 62(2), pp.782-790. | 3327 | 60.8 |
| 18 | Klein, S., Staring, M., Murphy, K., Viergever, M.A. and Pluim, J.P., 2009. Elastix: a toolbox for intensity-based medical image registration. *IEEE transactions on medical imaging*, 29(1), pp.196-205. | 1547 | 59.5 |
| 19 | Lemm, S., Blankertz, B., Dickhaus, T. and Müller, K.R., 2011. Introduction to machine learning for brain imaging. *Neuroimage*, 56(2), pp.387-399. | 322 | 58.2 |
| 20 | Potter, E. and Marwick, T.H., 2018. Assessment of left ventricular function by echocardiography: the case for routinely adding global longitudinal strain to ejection fraction. *JACC: Cardiovascular Imaging*, 11(2 Part 1), pp.260-274. | 57 | 56.0 |
| 21 | Boellaard, R., Delgado-Bolton, R., Oyen, W.J., Giammarile, F., Tatsch, K., Eschner, W., Verzijlbergen, F.J., Barrington, S.F., Pike, L.C., Weber, W.A. and Stroobants, S., 2015. FDG PET/CT: EANM procedure guidelines for tumour imaging: version 2.0. *European journal of nuclear medicine and molecular imaging*, 42(2), pp.328-354. | 682 | 55.8 |
| 22 | Winkler, A.M., Ridgway, G.R., Webster, M.A., Smith, S.M. and Nichols, T.E., 2014. Permutation inference for the general linear model. *Neuroimage*, 92, pp.381-397. | 1084 | 55.7 |
| 23 | McCollough, C.H., Leng, S., Yu, L., Cody, D.D., Boone, J.M. and McNitt-Gray, M.F., 2011. CT dose index and patient dose: they are not the same thing. *Radiology*, 259(2), pp.311-316. | 216 | 54.6 |
| 24 | MacMahon, H., Naidich, D.P., Goo, J.M., Lee, K.S., Leung, A.N., Mayo, J.R., Mehta, A.C., Ohno, Y., Powell, C.A., Prokop, M. and Rubin, G.D., 2017. Guidelines for management of incidental pulmonary nodules detected on CT images: from the Fleischner Society 2017. *Radiology*, 284(1), pp.228-243. | 397 | 54.1 |
| 25 | Wald, C., Russo, M.W., Heimbach, J.K., Hussain, H.K., Pomfret, E.A. and Bruix, J., 2013. New OPTN/UNOS policy for liver transplant allocation: standardization of liver imaging, diagnosis, classification, and reporting of hepatocellular carcinoma. | 190 | 53.8 |
| 26 | Tajbakhsh, N., Shin, J.Y., Gurudu, S.R., Hurst, R.T., Kendall, C.B., Gotway, M.B. and Liang, J., 2016. Convolutional neural networks for medical image analysis: Full training or fine tuning?. *IEEE transactions on medical imaging*, 35(5), pp.1299-1312. | 557 | 52.3 |
| 27 | Schauer, D.A. and Linton, O.W., 2009. NCRP report No. 160, ionizing radiation exposure of the population of the United States, medical exposure—are we doing less with more, and is there a role for health physicists?. *Health physics*, 97(1), pp.1-5. | 139 | 52.0 |
| 28 | Van Essen, D.C., Smith, S.M., Barch, D.M., Behrens, T.E., Yacoub, E., Ugurbil, K. and Wu-Minn HCP Consortium, 2013. The WU-Minn human connectome project: an overview. *Neuroimage*, 80, pp.62-79. | 1168 | 51.3 |
| 29 | McDonald, R.J., McDonald, J.S., Kallmes, D.F., Jentoft, M.E., Murray, D.L., Thielen, K.R., Williamson, E.E. and Eckel, L.J., 2015. Intracranial gadolinium deposition after contrast-enhanced MR imaging. *Radiology*, 275(3), pp.772-782. | 619 | 50.7 |
| 30 | Menze, B.H., Jakab, A., Bauer, S., Kalpathy-Cramer, J., Farahani, K., Kirby, J., Burren, Y., Porz, N., Slotboom, J., Wiest, R. and Lanczi, L., 2014. The multimodal brain tumor image segmentation benchmark (BRATS). *IEEE transactions on medical imaging*, 34(10), pp.1993-2024. | 706 | 50.6 |
| 31 | Van Dijk, K.R., Sabuncu, M.R. and Buckner, R.L., 2012. The influence of head motion on intrinsic functional connectivity MRI. *Neuroimage*, 59(1), pp.431-438. | 1351 | 50.1 |
| 32 | Wang, G., Ye, J.C., Mueller, K. and Fessler, J.A., 2018. Image reconstruction is a new frontier of machine learning. *IEEE transactions on medical imaging*, 37(6), pp.1289-1296. | 42 | 49.4 |
| 33 | Reeder, S.B., Hu, H.H. and Sirlin, C.B., 2012. Proton density fat-fraction: a standardized MR-based biomarker of tissue fat concentration. *Journal of magnetic resonance imaging: JMIR*, 36(5), p.1011. | 184 | 49.3 |
| 34 | Kanal, E. and Tweedle, M.F., 2015. Residual or retained gadolinium: practical implications for radiologists and our patients. *Radiology*, 275(3), pp.630-634. | 124 | 48.0 |
| 35 | Power, J.D., Mitra, A., Laumann, T.O., Snyder, A.Z., Schlaggar, B.L. and Petersen, S.E., 2014. Methods to detect, characterize, and remove motion artifact in resting state fMRI. *Neuroimage*, 84, pp.320-341. | 925 | 47.5 |
| 36 | Andersson, J.L. and Sotiropoulos, S.N., 2016. An integrated approach to correction for off-resonance effects and subject movement in diffusion MR imaging. *Neuroimage*, 125, pp.1063-1078. | 538 | 45.6 |
| 37 | Silva, A.C., Morse, B.G., Hara, A.K., Paden, R.G., Hongo, N. and Pavlicek, W., 2011. Dual-energy (spectral) CT: applications in abdominal imaging. *Radiographics*, 31(4), pp.1031-1046. | 180 | 45.5 |
| 38 | Vallières, M., Zwanenburg, A., Badic, B., Le Rest, C.C., Visvikis, D. and Hatt, M., 2018. Responsible radiomics research for faster clinical translation. | 45 | 44.2 |
| 39 | Eiber, 2016. "Evaluation of Hybrid Ga-68-PSMA Ligand PET/CT in 248 Patients with Biochemical Recurrence After Radical Prostatectomy"(vol 56, pg 668, 2015). *Journal of nuclear medicine*, 57(8), pp.1325-1325. | 538 | 44.0 |
| 40 | Tustison, N.J., Avants, B.B., Cook, P.A., Zheng, Y., Egan, A., Yushkevich, P.A. and Gee, J.C., 2010. N4ITK: improved N3 bias correction. *IEEE transactions on medical imaging*, 29(6), pp.1310-1320. | 1131 | 43.5 |
| 41 | Glasser, M.F., Sotiropoulos, S.N., Wilson, J.A., Coalson, T.S., Fischl, B., Andersson, J.L., Xu, J., Jbabdi, S., Webster, M., Polimeni, J.R. and Van Essen, D.C., 2013. The minimal preprocessing pipelines for the Human Connectome Project. *Neuroimage*, 80, pp.105-124. | 987 | 43.4 |
| 42 | Johnson, N.P., Kirkeeide, R.L. and Gould, K.L., 2012. Is discordance of coronary flow reserve and fractional flow reserve due to methodology or clinically relevant coronary pathophysiology? *JACC: Cardiovascular Imaging*, 5(2), pp.193-202. | 162 | 43.1 |
| 43 | Pereira, S., Pinto, A., Alves, V. and Silva, C.A., 2016. Brain tumor segmentation using convolutional neural networks in MRI images. *IEEE transactions on medical imaging*, 35(5), pp.1240-1251. | 458 | 43.0 |
| 44 | Afshar-Oromieh, A., Avtzi, E., Giesel, F.L., Holland-Letz, T., Linhart, H.G., Eder, M., Eisenhut, M., Boxler, S., Hadaschik, B.A., Kratochwil, C. and Weichert, W., 2015. The diagnostic value of PET/CT imaging with the 68 Ga-labelled PSMA ligand HBED-CC in the diagnosis of recurrent prostate cancer. *European journal of nuclear medicine and molecular imaging*, 42(2), pp.197-209. | 498 | 40.8 |
| 45 | Pibarot, P., Hahn, R.T., Weissman, N.J. and Monaghan, M.J., 2015. Assessment of paravalvular regurgitation following TAVR: a proposal of unifying grading scheme. *JACC: Cardiovascular Imaging*, 8(3), pp.340-360. | 106 | 40.5 |
| 46 | Le Bihan, D., 2013. Apparent diffusion coefficient and beyond: what diffusion MR imaging can tell us about tissue structure. | 143 | 40.5 |
| 47 | Hutchison, R.M., Womelsdorf, T., Allen, E.A., Bandettini, P.A., Calhoun, V.D., Corbetta, M., Della Penna, S., Duyn, J.H., Glover, G.H., Gonzalez-Castillo, J. and Handwerker, D.A., 2013. Dynamic functional connectivity: promise, issues, and interpretations. *Neuroimage*, 80, pp.360-378. | 920 | 40.4 |
| 48 | Grayburn, P.A., Sannino, A. and Packer, M., 2019. Proportionate and disproportionate functional mitral regurgitation: a new conceptual framework that reconciles the results of the MITRA-FR and COAPT trials. *JACC: Cardiovascular Imaging*, 12(2), pp.353-362. | 68 | 39.7 |
| 49 | Fleischmann, D. and Boas, F.E., 2011. Computed tomography—old ideas and new technology. | 153 | 38.7 |
| 50 | Avants, B.B., Tustison, N.J., Song, G., Cook, P.A., Klein, A. and Gee, J.C., 2011. A reproducible evaluation of ANTs similarity metric performance in brain image registration. *Neuroimage*, 54(3), pp.2033-2044. | 1130 | 38.1 |
| 51 | Blanke, P., Naoum, C., Dvir, D., Bapat, V., Ong, K., Muller, D., Cheung, A., Ye, J., Min, J.K., Piazza, N. and Theriault-Lauzier, P., 2017. Predicting LVOT obstruction in transcatheter mitral valve implantation: concept of the neo-LVOT. *JACC: Cardiovascular Imaging*, 10(4), pp.482-485. | 65 | 37.8 |
| 52 | Jacques, S.L., 2013. Optical properties of biological tissues: a review. *Physics in Medicine & Biology*, 58(11), p.R37. | 1317 | 37.2 |
| 53 | Murphy, K., Birn, R.M., Handwerker, D.A., Jones, T.B. and Bandettini, P.A., 2009. The impact of global signal regression on resting state correlations: are anti-correlated networks introduced? *Neuroimage*, 44(3), pp.893-905. | 1353 | 37.0 |
| 54 | Kanda, T., Ishii, K., Kawaguchi, H., Kitajima, K. and Takenaka, D., 2014. High signal intensity in the dentate nucleus and globus pallidus on unenhanced T1-weighted MR images: relationship with increasing cumulative dose of a gadolinium-based contrast material. *Radiology*, 270(3), pp.834-841. | 541 | 36.3 |
| 55 | Patenaude, B., Smith, S.M., Kennedy, D.N. and Jenkinson, M., 2011. A Bayesian model of shape and appearance for subcortical brain segmentation. *Neuroimage*, 56(3), pp.907-922. | 1065 | 35.9 |
| 56 | Tessler, F.N., Middleton, W.D., Grant, E.G., Hoang, J.K., Berland, L.L., Teehey, S.A., Cronan, J.J., Beland, M.D., Desser, T.S., Frates, M.C. and Hammers, L.W., 2017. ACR thyroid imaging, reporting and data system (TI-RADS): white paper of the ACR TI-RADS committee. *Journal of the American college of radiology*, 14(5), pp.587-595. | 259 | 35.3 |
| 57 | Kuo, M.D. and Jamshidi, N., 2014. Behind the numbers: decoding molecular phenotypes with radiogenomics—guiding principles and technical considerations. *Radiology*, 270(2), pp.320-325. | 90 | 34.6 |
| 58 | Lakhani, P. and Sundaram, B., 2017. Deep learning at chest radiography: automated classification of pulmonary tuberculosis by using convolutional neural networks. *Radiology*, 284(2), pp.574-582. | 251 | 34.2 |
| 59 | Madabhushi, A. and Lee, G., 2016. Image analysis and machine learning in digital pathology: Challenges and opportunities. | 124 | 34.2 |
| 60 | Afshar-Oromieh, A., Zechmann, C.M., Malcher, A., Eder, M., Eisenhut, M., Linhart, H.G., Holland-Letz, T., Hadaschik, B.A., Giesel, F.L., Debus, J. and Haberkorn, U., 2014. Comparison of PET imaging with a 68 Ga-labelled PSMA ligand and 18 F-choline-based PET/CT for the diagnosis of recurrent prostate cancer. *European journal of nuclear medicine and molecular imaging*, 41(1), pp.11-20. | 506 | 33.9 |
| 61 | Bamber, J., Cosgrove, D., Dietrich, C.F., Fromageau, J., Bojunga, J., Calliada, F., Cantisani, V., Correias, J.M., D'Onofrio, M., Drakonaki, E.E. and Fink, M., 2013. EFSUMB guidelines and recommendations on the clinical use of ultrasound elastography. Part 1: Basic principles and technology. *Ultraschall in der Medizin-European Journal of Ultrasound*, 34(02), pp.169-184. | 523 | 33.6 |
| 62 | Cerqueira, M.D., Allman, K.C., Ficaro, E.P., Hansen, C.L., Nichols, K.J., Thompson, R.C., Van Decker, W.A. and Yakovlevitch, M., 2010. Recommendations for reducing radiation exposure in myocardial perfusion imaging. *Journal of nuclear cardiology*, 17(4), pp.709-718. | 175 | 32.9 |
| 63 | Fischl, B., 2012. FreeSurfer. *Neuroimage*, 62(2), pp.774-781. | 1797 | 32.8 |
| 64 | Klein, A., Andersson, J., Ardekani, B.A., Ashburner, J., Avants, B., Chiang, M.C., Christensen, G.E., Collins, D.L., Gee, J., Hellier, P. and Song, J.H., 2009. Evaluation of 14 nonlinear deformation algorithms applied to human brain MRI registration. *Neuroimage*, 46(3), pp.786-802. | 1188 | 32.5 |
| 65 | Kanda, T., Fukusato, T., Matsuda, M., Toyoda, K., Oba, H., Kotoku, J.I., Haruyama, T., Kitajima, K. and Furui, S., 2015. Gadolinium-based contrast agent accumulates in the brain even in subjects without severe renal dysfunction: evaluation of autopsy brain specimens with inductively coupled plasma mass spectroscopy. *Radiology*, 276(1), pp.228-232. | 393 | 32.2 |
| 66 | Reuter, M., Schmansky, N.J., Rosas, H.D. and Fischl, B., 2012. Within-subject template estimation for unbiased longitudinal image analysis. *Neuroimage*, 61(4), pp.1402-1418. | 851 | 31.6 |
| 67 | Kabasakal, E.D.L. and Kanmaz, M.H.B., 2014. 68 Ga-PSMA PET/CT imaging of metastatic clear cell renal cell carcinoma. *Eur. J. Nucl. Med. Mol. Imaging*, 41, pp.1461-1462. | 82 | 31.5 |
| 68 | Jack Jr, C.R., 2012. Alzheimer disease: new concepts on its neurobiology and the clinical role imaging will play. *Radiology*, 263(2), pp.344-361. | 117 | 31.3 |
| 69 | Woolrich, M.W., Jbabdi, S., Patenaude, B., Chappell, M., Makni, S., Behrens, T., Beckmann, C., Jenkinson, M. and Smith, S.M., 2009. Bayesian analysis of neuroimaging data in FSL. *Neuroimage*, 45(1), pp.S173-S186. | 1142 | 31.2 |
| 70 | Cosgrove, D., Piscaglia, F., Bamber, J., Bojunga, J., Correias, J.M., Gilja, O.H., Klausner, A.S., Sporea, I., Calliada, F., Cantisani, V. and D'Onofrio, M., 2013. EFSUMB guidelines and recommendations on the clinical use of ultrasound elastography. Part 2: Clinical applications. *Ultraschall in der Medizin-European Journal of Ultrasound*, 34(03), pp.238-253. | 482 | 31.0 |
| 71 | Yan, C.G., Cheung, B., Kelly, C., Colcombe, S., Craddock, R.C., Di Martino, A., Li, Q., Zuo, X.N., Castellanos, F.X. and Milham, M.P., 2013. A comprehensive assessment of regional variation in the impact of head micromovements on functional connectomics. *Neuroimage*, 76, pp.183-201. | 703 | 30.9 |
| 72 | Zhang, H., Schneider, T., Wheeler-Kingshott, C.A. and Alexander, D.C., 2012. NODDI: practical in vivo neurite orientation dispersion and density imaging of the human brain. *Neuroimage*, 61(4), pp.1000-1016. | 831 | 30.8 |
| 73 | International Commission on Non-Ionizing Radiation Protection, 2010. Guidelines for limiting exposure to time-varying electric and magnetic fields (1 Hz to 100 kHz). *Health physics*, 99(6), pp.818-836. | 582 | 30.2 |
| 74 | Walsh, L., Shore, R., Auvinen, A., Jung, T. and Wakeford, R., 2014. Risks from CT scans—what do recent studies tell us?. *Journal of radiological protection*, 34(1), p.E1. | 56 | 30.0 |
| 75 | Sotiras, A., Davatzikos, C. and Paragios, N., 2013. Deformable medical image registration: A survey. *IEEE transactions on medical imaging*, 32(7), pp.1153-1190. | 552 | 29.9 |
| 76 | Clark, K., Vendt, B., Smith, K., Freymann, J., Kirby, J., Koppel, P., Moore, S., Phillips, S., Maffitt, D., Pringle, M. and Tarbox, L., 2013. The Cancer Imaging Archive (TCIA): maintaining and operating a public information repository. *Journal of digital imaging*, 26(6), pp.1045-1057. | 527 | 29.7 |
| 77 | Wu, C.C., Maher, M.M. and Shepard, J.A.O., 2011. Complications of CT-guided percutaneous needle biopsy of the chest: prevention and management. *American Journal of Roentgenology*, 196(6), pp.W678-W682. | 117 | 29.6 |
| 78 | Smith, S.M., Miller, K.L., Salimi-Khorshidi, G., Webster, M., Beckmann, C.F., Nichols, T.E., Ramsey, J.D. and Woolrich, M.W., 2011. Network modelling methods for FMRI. *Neuroimage*, 54(2), pp.875-891. | 873 | 29.4 |
| 79 | Horn, A., Li, N., Dembek, T.A., Kappel, A., Boulay, C., Ewert, S., Tietze, A., Husch, A., Perera, T., Neumann, W.J. and Reiser, M., 2019. Lead-DBS v2: Towards a comprehensive pipeline for deep brain stimulation imaging. *Neuroimage*, 184, pp.293-316. | 50 | 29.3 |
| 80 | Piscaglia, F., Nolsøe, C., Dietrich, C.A., Cosgrove, D.O., Gilja, O.H., Nielsen, M.B., Albrecht, T., Barozzi, L., Bertolotto, M., Catalano, O. and Claudon, M., 2012. The EFSUMB Guidelines and Recommendations on the Clinical Practice of Contrast Enhanced Ultrasound (CEUS): update 2011 on non-hepatic applications. *Ultraschall in der Medizin-European Journal of Ultrasound*, 33(01), pp.33-59. | 506 | 29.3 |
| 81 | Boellaard, R., O'Doherty, M.J., Weber, W.A., Mottaghy, F.M., Lonsdale, M.N., Stroobants, S.G., Oyen, W.J., Kotzerke, J., Hoekstra, O.S., Pruim, J. and Marsden, P.K., 2010. FDG PET and PET/CT: EANM procedure guidelines for tumour PET imaging: version 1.0. *European journal of nuclear medicine and molecular imaging*, 37(1), p.181. | 730 | 29.2 |
| 82 | Afshar-Oromieh, A., Haberkorn, U., Eder, M., Eisenhut, M. and Zechmann, C.M., 2012. [68 Ga] Gallium-labelled PSMA ligand as superior PET tracer for the diagnosis of prostate cancer: comparison with 18 F-FECH. *European journal of nuclear medicine and molecular imaging*, 39(6), pp.1085-1086. | 109 | 29.2 |
| 83 | Zalesky, A. and Breakspear, M., 2015. Towards a statistical test for functional connectivity dynamics. *Neuroimage*, 114, pp.466-470. | 102 | 29.1 |
| 84 | Lamm, C., Decety, J. and Singer, T., 2011. Meta-analytic evidence for common and distinct neural networks associated with directly experienced pain and empathy for pain. *Neuroimage*, 54(3), pp.2492-2502. | 857 | 28.9 |
| 85 | Giesel, F.L., Knorr, K., Spohn, F., Will, L., Maurer, T., Flechsig, P., Neels, O., Schiller, K., Amaral, H., Weber, W.A. and Haberkorn, U., 2019. Detection efficacy of 18F-PSMA-1007 PET/CT in 251 | 42 | 28.9 |
|  | patients with biochemical recurrence of prostate cancer after radical prostatectomy. *Journal of Nuclear Medicine*, 60(3), pp.362-368. |  |  |
| 86 | Naidich, D.P., Bankier, A.A., MacMahon, H., Schaefer-Prokop, C.M., Pistoletti, M., Goo, J.M., Macchiarini, P., Crapo, J.D., Herold, C.J., Austin, J.H. and Travis, W.D., 2013. Recommendations for the management of subsolid pulmonary nodules detected at CT: a statement from the Fleischner Society. *Radiology*, 266(1), pp.304-317. | 510 | 28.8 |
| 87 | Anthimopoulos, M., Christodoulidis, S., Ebner, L., Christe, A. and Mougiakakou, S., 2016. Lung pattern classification for interstitial lung diseases using a deep convolutional neural network. *IEEE transactions on medical imaging*, 35(5), pp.1207-1216. | 306 | 28.7 |
| 88 | Darcourt, J., Booi, J., Tatsch, K., Varrone, A., Vander Borght, T., Kapucu, Ö.L., Nägren, K., Nobili, F., Walker, Z. and Van Laere, K., 2010. EANM procedure guidelines for brain neurotransmission SPECT using 123 I-labelled dopamine transporter ligands, version 2. *European journal of nuclear medicine and molecular imaging*, 37(2), pp.443-450. | 150 | 28.5 |
| 89 | Satterthwaite, T.D., Elliott, M.A., Gerraty, R.T., Ruparel, K., Loughhead, J., Calkins, M.E., Eickhoff, S.B., Hakonarson, H., Gur, R.C., Gur, R.E. and Wolf, D.H., 2013. An improved framework for confound regression and filtering for control of motion artifact in the preprocessing of resting-state functional connectivity data. *Neuroimage*, 64, pp.240-256. | 645 | 28.4 |
| 90 | Power, J.D., Barnes, K.A., Snyder, A.Z., Schlaggar, B.L. and Petersen, S.E., 2013. Steps toward optimizing motion artifact removal in functional connectivity MRI; a reply to Carp. *Neuroimage*, 76. | 173 | 28.3 |
| 91 | Bartra, O., McGuire, J.T. and Kable, J.W., 2013. The valuation system: a coordinate-based meta-analysis of BOLD fMRI experiments examining neural correlates of subjective value. *Neuroimage*, 76, pp.412-427. | 637 | 28.0 |
| 92 | Christ, A., Kainz, W., Hahn, E.G., Honegger, K., Zefferer, M., Neufeld, E., Rascher, W., Janka, R., Bautz, W., Chen, J. and Kiefer, B., 2009. The Virtual Family—development of surface-based anatomical models of two adults and two children for dosimetric simulations. *Physics in Medicine & Biology*, 55(2), p.N23. | 767 | 28.0 |
| 93 | Lammer, J., Malagari, K., Vogl, T., Pilleul, F., Denys, A., Watkinson, A., Pitton, M., Sergeant, G., Pfammatter, T., Terraz, S. and Benhamou, Y., 2010. Prospective randomized study of doxorubicin-eluting-bead embolization in the treatment of hepatocellular carcinoma: results of the PRECISION V study. *Cardiovascular and interventional radiology*, 33(1), pp.41-52. | 737 | 27.9 |
| 94 | Setio, A.A.A., Ciompi, F., Litjens, G., Gerke, P., Jacobs, C., Van Riel, S.J., Wille, M.M.W., Naqibullah, M., Sánchez, C.I. and van Ginneken, B., 2016. Pulmonary nodule detection in CT images: false positive reduction using multi-view convolutional networks. *IEEE transactions on medical imaging*, 35(5), pp.1160-1169. | 296 | 27.8 |
| 95 | Kumar, V., Gu, Y., Basu, S., Berglund, A., Eschrich, S.A., Schabath, M.B., Forster, K., Aerts, H.J., Dekker, A., Fenstermacher, D. and Goldgof, D.B., 2012. Radiomics: the process and the challenges. *Magnetic resonance imaging*, 30(9), pp.1234-1248. | 557 | 27.7 |
| 96 | Mazurowski, M.A., 2015. Radiogenomics: what it is and why it is important. *Journal of the American College of Radiology*, 12(8), pp.862-866. | 71 | 27.5 |
| 97 | Greve, D.N. and Fischl, B., 2009. Accurate and robust brain image alignment using boundary-based registration. *Neuroimage*, 48(1), pp.63-72. | 1002 | 27.4 |
| 98 | Villemagne, V.L., Klunk, W.E., Mathis, C.A., Rowe, C.C., Brooks, D.J., Hyman, B.T., Ikonomic, M.D., Ishii, K., Jack, C.R., Jagust, W.J. and Johnson, K.A., 2012. Aβ Imaging: feasible, pertinent, and vital to progress in Alzheimer's disease. | 102 | 27.3 |
| 99 | Marinescu, M.A., Löffler, A.I., Ouellette, M., Smith, L., Kramer, C.M. and Bourque, J.M., 2015. Coronary microvascular dysfunction, microvascular angina, and treatment strategies. *JACC: Cardiovascular Imaging*, 8(2), pp.210-220. | 71 | 27.1 |
| 100 | Kooi, T., Litjens, G., Van Ginneken, B., Gubern-Mérida, A., Sánchez, C.I., Mann, R., den Heeten, A. and Karssemeijer, N., 2017. Large scale deep learning for computer aided detection of mammographic lesions. *Medical image analysis*, 35, pp.303-312. | 215 | 27.0 |

**Table 3.** Number of articles per list according to year of publication.

| Year of publication | No. of articles in highest-cited list | No. of articles in highest-CNCI list |
| --- | --- | --- |
| 2009 | 22 | 7 |
| 2010 | 22 | 10 |
| 2011 | 8 | 9 |
| 2012 | 18 | 15 |
| 2013 | 11 | 16 |
| 2014 | 5 | 7 |
| 2015 | 6 | 12 |
| 2016 | 5 | 10 |
| 2017 | 3 | 8 |
| 2018 | 0 | 3 |
| 2020 | 0 | 3 |

**Figure 1.**
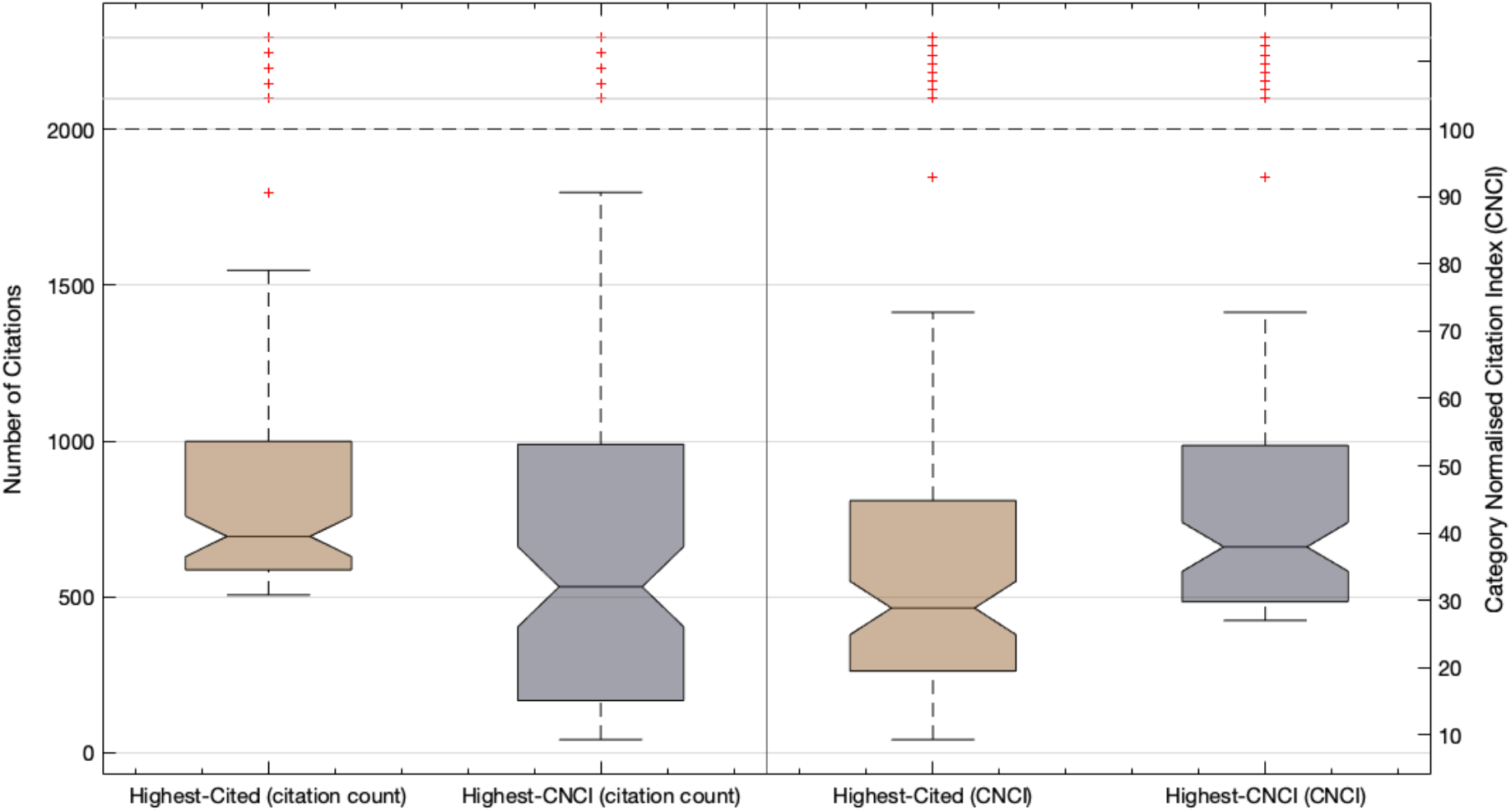
Box plot illustrating distribution of number citations (left axis) and category normalised impact factor (right axis) for the highest-cited (grey) and highest-impact (brown) article lists

The highest-cited works appeared across 23 different journals while the 100 highest-CNCI across 24 (Table 4). The journal with greatest number of highest-cited and highest-CNCI articles was *NeuroImage* with 51 and 30 respectively. The next highest contributor in the most-cited list was *IEEE Transactions on Medical Imaging* (n = 6), *Human Brain Mapping* (n = 5), and *Magnetic Resonance in Imaging* (n = 5). Within the highest-CNCI list different journals showed higher contributors of articles with *Radiology* (n = 13), *IEEE Transactions on Medical Imaging* (n = 11), and *European Journal of Nuclear Medicine and Molecular Imaging* (n = 8) contributing the most.

**Table 4.** Number of articles per list according to publishing journal.

| **Journal** | **No. of articles in highest-cited list** | **No. of articles in highest-CNCI list** |
| --- | --- | --- |
| *NeuroImage* | 51 | 30 |
| *IEEE Transactions on Medical Imaging* | 6 | 11 |
| *Human Brain Mapping* | 5 | 0 |
| *Magnetic Resonance in Medicine* | 5 | 0 |
| *Medical Image Analysis* | 4 | 5 |
| *Radiology* | 4 | 13 |
| *European Journal of Nuclear Medicine* | 3 | 8 |
| *European Journal of Echocardiography* | 2 | 0 |
| *Journal of Biomedical Optics* | 2 | 0 |
| *Magnetic Resonance Imaging* | 2 | 2 |
| *Physics in Medicine and Biology* | 2 | 2 |
| *Ultraschall In Der Medizin* | 2 | 3 |
| *American Journal of Roentgenology* | 1 | 1 |
| *Cardiovascular and Interventional Radiology* | 1 | 1 |
| *European Heart Journal – Cardiology* | 1 | 1 |
| *European Radiology* | 1 | 2 |
| *Health Physics* | 1 | 2 |
| *International Journal of Computer Assisted Radiology and Surgery* | 1 | 0 |
| *International Journal of Radiation Oncology – Biology – Physics* | 1 | 0 |
| *Journal of Digital Imaging* | 1 | 1 |
| *Journal of Nuclear Medicine* | 1 | 4 |
| *Journal of the American Society of Echocardiography* | 1 | 0 |
| *Medical Physics* | 1 | 0 |
| *NMR in Biomedicine* | 1 | 0 |
| *JACC – Cardiovascular Imaging* | 0 | 6 |
| *Journal of Magnetic Resonance* | 0 | 1 |
| *Journal of Nuclear Cardiology* | 0 | 1 |
| *Journal of Radiological Protection* | 0 | 1 |
| *Journal of the American College of Radiology* | 0 | 2 |
| *Radiation Protection Dosimetry* | 0 | 1 |
| *Radiographics* | 0 | 1 |
| *Ultrasound in Obstetrics and Gynaecology* | 0 | 1 |

The most frequent referenced subspeciality and modality was common to both the highest-citation and highest-CNCI list. These were neuroradiology (68 and 41 articles) and magnetic resonance imaging (64 and 40 articles), nothing these are not independent variables with MRI the primarily modality in neuroradiological imaging. Sub-analysis revealed that of the 68 and 41 neuroradiology articles within each list, 55% and 58% of each pertained to development or evaluation of computer science/mathematical/image processing techniques on neuroradiological data rather than primary clinical radiological research.

Articles conforming to ‘miscellaneous’ (15 & 21 articles) and imaging ‘safety’ (5, 6 articles respectively) were the next most common radiological domain studied for both list. The highest-CNCI articles demonstrated a greater variance in modalities studied in comparison to the highest-citation list. In particular, a greater number of studies with primary modality of nuclear medicine and CT were present in the highest-CNCI list. A breakdown of subspecialty and modality is given in Table 5.

**Table 5.** Number of articles according to radiological subspecialty and primary modality

| ***Radiological Subspecialty*** | **No. of articles in highest-cited list** | **No. of articles in highest-CNCI list** |
| --- | --- | --- |
| Neuroradiology | 68 | 41 |
| Cardiac | 4 | 7 |
| Nuclear Imaging | 4 | 7 |
| Interventional | 1 | 2 |
| Genitourinary | 1 | 5 |
| Respiratory | 1 | 6 |
| Abdominal | 0 | 2 |
| Breast | 0 | 1 |
| Mixed | 1 | 2 |
| Safety | 5 | 6 |
| Miscellaneous | 15 | 21 |
| ***Primary Modality*** |  |  |
| MR Imaging | 64 | 40 |
| US | 7 | 7 |
| CT | 4 | 12 |
| Nuclear Imaging | 4 | 13 |
| Interventional | 1 | 1 |
| Breast imaging | 0 | 1 |
| Radiography | 0 | 1 |
| Mixed modality | 8 | 9 |
| Other | 12 | 16 |

## 4.0 Discussion

Within this study the first, to the authors knowledge, bibliometric analysis of radiological articles based on highest CNCI was performed. This was done alongside an analysis of the 100 highest raw citation count articles for the past decade of radiological literature. Similar bibliometric studies have been performed in the past, including analysis on the highest cited medical imaging literature ever published (7,10,11,13,14). However, with the ever-growing volume of academic publications, contemporaneous re-evaluation of emerging trends and topics, with alternative metrics, is beneficial to inform various stakeholders. These stakeholders include (but are not limited to) practicing and trainee radiologists, physicists/mathematicians, computer scientists, radiographers, and nonradiologist clinicians.

The decade of 2009 – 2019 highest-cited articles had characteristics similar to previous studies (7,8,11). In particular neuroradiology and magnetic resonance imaging were highly prevalent topics of research. An explanation for this is proposed by Brinkiji et al. in which they discuss how there is a variety of stakeholders from various research fields publishing on this topic resulting in a skewed representation of neuroradiologically inclined research (11). The populist nature of these fields results in an inherent bias in the number of citations a publication with garner. The analysis presented here of the temporal course of the highest-cited works show a predilection for older articles, median year of 2011, with most recent work appearing in the list published in 2017. This highlights another bias in relying on raw citation count as a metric of academic performance. In particular, older articles have a greater amount of time to accrue citations when compared to contemporary works. The CNCI of these articles show a broad range with a median of CNCI of 28.9. Whilst these articles show a lower median/average CNCI than the highest-CNCI list it is important to note that even the article with the lowest CNCI (9.3) still garners 9.3x as many citations as expected for an article of same type and same year of publication.

The highest-CNCI list, in comparison, shows greater diversity in its citation range, subspeciality referenced and imaging modality and differences in temporal trends. Objectively, the lists have a low similarity score with Jaccard index of 33% resenting significant differences in their contents. Whilst the most dominant subspeciality and modality remains neuroradiology and magnetic resonance imaging they it is a smaller comparison to the highest cited list. With respect to subspeciality, there is a greater spread of research areas contained within the list. In particular, articles related to nuclear imaging, respiratory system, and genitourinary systems are more abundant. Further, the highest-CNCI articles have a greater diversity in other imaging modalities, including a small number of articles based on conventional radiography and breast imaging. The emergence of these studies illustrate the utility of normalised bibliometric indices, here the CNCI. In particular, whilst neuroradiology and MR imaging may have the greatest volume of researchers and overlap with computer science, when normalised to their domain of study other fields can be more fairly compared. In addition, temporal citation bias is removed by indexing its performance to the year of publication. This is evidenced by the even distribution and spread of article publication year in the highest-CNCI articles. As an example, the article “Dual-Energy (Spectral) CT: Applications in Abdominal Imaging” appears in the highest-CNCI list whose subspecialty and modality is not represented well within the highest-cited list. However, spectral imaging is an emerging field of radiology with a great deal of potential and interest to the radiological community at large (15,16,16,17). As such the inherent value in understanding and evaluating articles based on alternative bibliometric values is apparent. One possibility is that by using normalised bibliometric analysis ‘classic’ or ‘seminal’ works can be identified or predicted prospectively.

There are limitations of this study. Citation analysis is not a precise science, nor is normalisation with respect to category normalised citation index. Both metrics of interest are inherently biased by self-citation and the so-called ‘Bradfords’ Law. This law describing the positive feedback mechanism of publishing and referencing from a small variety of journals or authors (18). Further, an articles CNCI may be artificially raised by including it into a category that does not typically garner many publications and rewards recent publications that have been cited. Those with low total citation counts and high CNCI need to be carefully considered as biased. As an example, the rank #86 article in the highest-impact list has 42 citations, where one would expect only 1.5 citations by now. However, given it has only recently been published it is inappropriate to evaluate the impact of this article purely on its CNCI value as the normalisation process requires an appropriate passage of time. Another limitation is the use of a single database, restricted to journals within the WOS “Radiology, Nuclear Medicine & Medical Imaging” SCIE and ESCI databases. Therefore, any articles that are radiologically based but published in journals outside those within the WOS database would not be included. This excludes radiology articles that may have been published in journals such as *Nature* and *Science*, for example. Further, if analysis were undertaken on different databases (e.g. Google Scholar, Scopus, etc) the highest-citation results would be expected to differ (19,20). However, the effect of differing databases on CNCI is not clear.

To mitigate and/or review the potential impact of these various limitations future researchers are encouraged to conduct a comparative analyses across all available databases, especially in regards to normalised citation rates. Further, to not exclude high profile radiological articles being excluded, more exhaustive searches for literature could be performed.

## 5.0 Conclusions

In summary, to the authors knowledge the first citation-normalised bibliometric analysis of radiological literature has been performed. This was done alongside a conventional raw citation count analysis over the years 2009 – 2019 in order to investigate the trends and characteristics of the highest-cited and highest-CNCI articles. The results illustrate the ongoing academic output the field of neuroradiology and use magnetic resonance imaging has. The CNCI analysis produced a list distinctly different from the highest-cited works with a more varied distribution of research subspecialty and imaging modality. This may better represent the current research landscape of radiological research and guide future bibliometric analyses.

## Data Availability

All data is available from lead author upon request.

## 7.0 Acknowledgements

The authors thank the input from Dr. Christopher Erian for useful discussion and guidance.

## Abbreviations

CNCI: Category Normalised Citation Index
WOS: Web of Science
SCIE: Science Citation Index Expanded
ESCI: Emerging Source Citation Index
MR: Magnetic Resonance

